## supplement for "The use of newborn foot length to identify low birth weight and preterm babies in Papua New Guinea: A diagnostic accuracy study"

### S1 Checklist. STARD checklist

|  | **Section & Topic** | **No** | **Item** | **Reported on page #** |
| --- | --- | --- | --- | --- |
|  | **TITLE OR ABSTRACT** |  |  | **1-2** |
|  |  | **1** | Identification as a study of diagnostic accuracy using at least one measure of accuracy  (such as sensitivity, specificity, predictive values, or AUC) | Title, p1 and Abstract p2 |
|  | **ABSTRACT** |  |  | ***2*** |
|  |  | **2** | Structured summary of study design, methods, results, and conclusions  (for specific guidance, see STARD for Abstracts) | 2 |
|  | **INTRODUCTION** |  |  | ***3*** |
|  |  | **3** | Scientific and clinical background, including the intended use and clinical role of the index test | 3 |
|  |  | **4** | Study objectives and hypotheses | 4 |
|  | **METHODS** |  |  | ***4-8*** |
|  | *Study design* | **5** | Whether data collection was planned before the index test and reference standard  were performed (prospective study) or after (retrospective study) | 4 |
|  | *Participants* | **6** | Eligibility criteria | 4 |
|  |  | **7** | On what basis potentially eligible participants were identified  (such as symptoms, results from previous tests, inclusion in registry) | 4 |
|  |  | **8** | Where and when potentially eligible participants were identified (setting, location and dates) | 4 |
|  |  | **9** | Whether participants formed a consecutive, random or convenience series | 4 |
|  | *Test methods* | **10a** | Index test, in sufficient detail to allow replication | 5 |
|  |  | **10b** | Reference standard, in sufficient detail to allow replication | 5 |
|  |  | **11** | Rationale for choosing the reference standard (if alternatives exist) | 3, 5 |
|  |  | **12a** | Definition of and rationale for test positivity cut-offs or result categories  of the index test, distinguishing pre-specified from exploratory | 7 |
|  |  | **12b** | Definition of and rationale for test positivity cut-offs or result categories  of the reference standard, distinguishing pre-specified from exploratory | 5 |
|  |  | **13a** | Whether clinical information and reference standard results were available  to the performers/readers of the index test | 6 |
|  |  | **13b** | Whether clinical information and index test results were available  to the assessors of the reference standard | 6 |
|  | *Analysis* | **14** | Methods for estimating or comparing measures of diagnostic accuracy | 7 |
|  |  | **15** | How indeterminate index test or reference standard results were handled | No missing data |
|  |  | **16** | How missing data on the index test and reference standard were handled | No missing data |
|  |  | **17** | Any analyses of variability in diagnostic accuracy, distinguishing pre-specified from exploratory | 7 |
|  |  | **18** | Intended sample size and how it was determined | 7-8 |
|  | **RESULTS** |  |  | ***8-13*** |
|  | *Participants* | **19** | Flow of participants, using a diagram | 8. Fig 2 |
|  |  | **20** | Baseline demographic and clinical characteristics of participants | 8, Table 1 (mothers);  10, Table 2 (babies) |
|  |  | **21a** | Distribution of severity of disease in those with the target condition | S1 Fig (LBW); Fig 3 (LBW and PTB) |
|  |  | **21b** | Distribution of alternative diagnoses in those without the target condition | Not applicable |
|  |  | **22** | Time interval and any clinical interventions between index test and reference standard | No interventions |
|  | *Test results* | **23** | Cross tabulation of the index test results (or their distribution)  by the results of the reference standard | Fig 4; Table 3; S1 Table, S2 Table |
|  |  | **24** | Estimates of diagnostic accuracy and their precision (such as 95% confidence intervals) | 11-12; Table 3; S1 Table, S2 Table |
|  |  | **25** | Any adverse events from performing the index test or the reference standard | 11 |
|  | **DISCUSSION** |  |  | ***13-16*** |
|  |  | **26** | Study limitations, including sources of potential bias, statistical uncertainty, and generalisability | 14 |
|  |  | **27** | Implications for practice, including the intended use and clinical role of the index test | 16 |
|  | **OTHER INFORMATION** |  |  |  |
|  |  | **28** | Registration number and name of registry | WANTAIM trial ISRCTN37134032 |
|  |  | **29** | Where the full study protocol can be accessed | Ethics committee submission |
|  |  | **30** | Sources of funding and other support; role of funders | 17-18 |

### S1 Fig. Distribution of average birth weight (n=342)

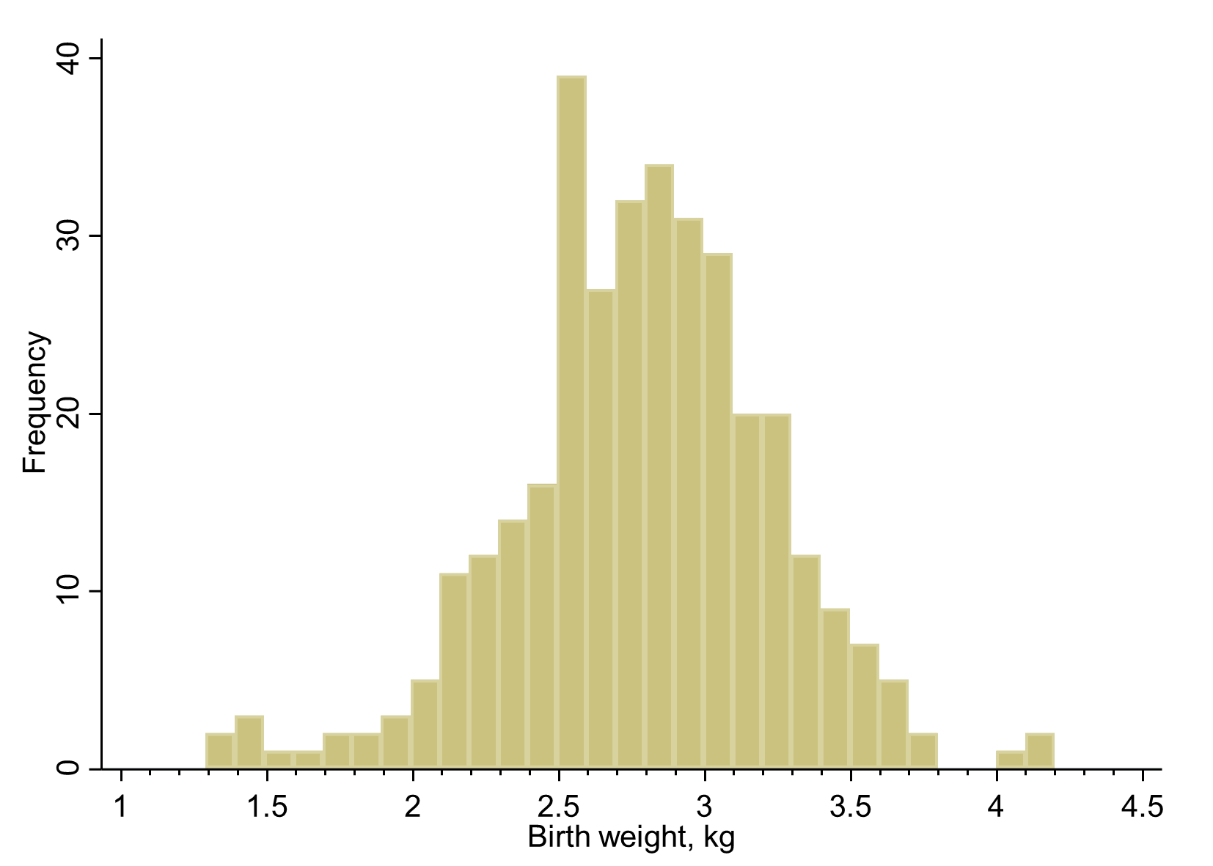

### S2 Fig. Distribution of average foot length measurements by A) clinical researcher (n=342), B) volunteer health care worker (n=123)

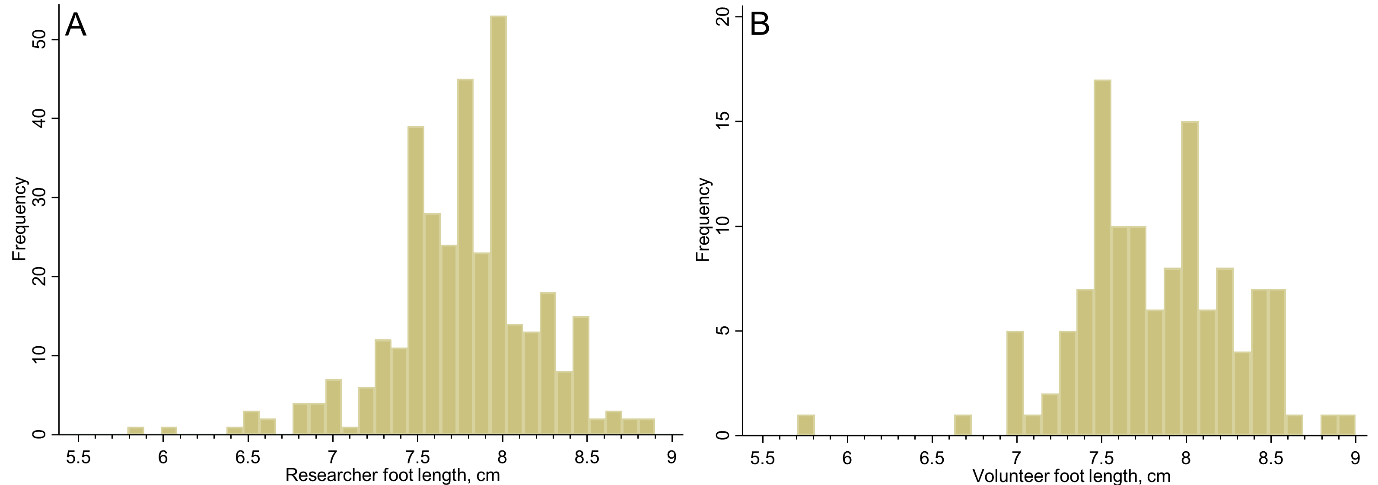

### S1 Table. Receiver operating curve statistics for accuracy of newborn foot length to classify low birth weight, 1 mm increments (N=342)

| **Low birth weight (<2.50 kg vs ≥2.50 kg)** | **True positive** | **False negative** | **False positive** | **True negative** | **Sensitivity, % (95% CI)** | **Specificity, % (95% CI)** | **Positive PV, % (95% CI)** | **Negative PV, % (95% CI)** | **Positive LHR, % (95% CI)** | **Negative LHR, % (95% CI)** | **AUC, % (95% CI)** | **Total, N** |
| --- | --- | --- | --- | --- | --- | --- | --- | --- | --- | --- | --- | --- |
| Researcher, overall |  |  |  |  |  |  |  |  |  |  | 87.0 (82.8-90.2) |  |
| Foot length (<5.8 cm) | 0 | 72 | 0 | 270 | 0.0 (0.0-5.1) | 100.0 (98.6-100.0) | .. | 78.9 (74.3-82.9) | .. | 1.0 (1.0-1.0) | 50.0 (44.6-55.4) | 342 |
| Foot length (<5.9 cm) | 1 | 71 | 0 | 270 | 1.4 (0.2-7.5) | 100.0 (98.6-100.0) | 100.0 (20.7-100.0) | 79.2 (74.6-83.2) | .. | 1.0 (1.0-1.0) | 50.7 (45.2-56.0) | 342 |
| Foot length (<6 cm) | 1 | 71 | 0 | 270 | 1.4 (0.2-7.5) | 100.0 (98.6-100.0) | 100.0 (20.7-100.0) | 79.2 (74.6-83.2) | .. | 1.0 (1.0-1.0) | 50.7 (45.2-56.0) | 342 |
| Foot length (<6.1 cm) | 2 | 70 | 0 | 270 | 2.8 (0.8-9.6) | 100.0 (98.6-100.0) | 100.0 (34.2-100.0) | 79.4 (74.8-83.4) | .. | 1.0 (0.9-1.0) | 51.4 (46.0-56.9) | 342 |
| Foot length (<6.2 cm) | 2 | 70 | 0 | 270 | 2.8 (0.8-9.6) | 100.0 (98.6-100.0) | 100.0 (34.2-100.0) | 79.4 (74.8-83.4) | .. | 1.0 (0.9-1.0) | 51.4 (46.0-56.9) | 342 |
| Foot length (<6.3 cm) | 2 | 70 | 0 | 270 | 2.8 (0.8-9.6) | 100.0 (98.6-100.0) | 100.0 (34.2-100.0) | 79.4 (74.8-83.4) | .. | 1.0 (0.9-1.0) | 51.4 (46.0-56.9) | 342 |
| Foot length (<6.4 cm) | 2 | 70 | 0 | 270 | 2.8 (0.8-9.6) | 100.0 (98.6-100.0) | 100.0 (34.2-100.0) | 79.4 (74.8-83.4) | .. | 1.0 (0.9-1.0) | 51.4 (46.0-56.9) | 342 |
| Foot length (<6.5 cm) | 3 | 69 | 0 | 270 | 4.2 (1.4-11.5) | 100.0 (98.6-100.0) | 100.0 (43.8-100.0) | 79.6 (75.0-83.6) | .. | 1.0 (0.9-1.0) | 52.1 (46.6-57.4) | 342 |
| Foot length (<6.6 cm) | 7 | 65 | 0 | 270 | 9.7 (4.8-18.7) | 100.0 (98.6-100.0) | 100.0 (64.6-100.0) | 80.6 (76.0-84.5) | .. | 0.9 (0.8-1.0) | 54.9 (49.5-60.3) | 342 |
| Foot length (<6.7 cm) | 7 | 65 | 0 | 270 | 9.7 (4.8-18.7) | 100.0 (98.6-100.0) | 100.0 (64.6-100.0) | 80.6 (76.0-84.5) | .. | 0.9 (0.8-1.0) | 54.9 (49.5-60.3) | 342 |
| Foot length (<6.8 cm) | 7 | 65 | 0 | 270 | 9.7 (4.8-18.7) | 100.0 (98.6-100.0) | 100.0 (64.6-100.0) | 80.6 (76.0-84.5) | .. | 0.9 (0.8-1.0) | 54.9 (49.5-60.3) | 342 |
| Foot length (<6.9 cm) | 11 | 61 | 0 | 270 | 15.3 (8.8-25.3) | 100.0 (98.6-100.0) | 100.0 (74.1-100.0) | 81.6 (77.0-85.4) | .. | 0.8 (0.8-0.9) | 57.6 (52.2-62.9) | 342 |
| Foot length (<7 cm) | 15 | 57 | 0 | 270 | 20.8 (13.1-31.6) | 100.0 (98.6-100.0) | 100.0 (79.6-100.0) | 82.6 (78.1-86.3) | .. | 0.8 (0.7-0.9) | 60.4 (55.1-65.7) | 342 |
| Foot length (<7.1 cm) | 22 | 50 | 2 | 268 | 30.6 (21.1-42.0) | 99.3 (97.3-99.8) | 91.7 (74.2-97.7) | 84.3 (79.9-87.9) | 41.2 (9.9-171.3) | 0.7 (0.6-0.8) | 64.9 (59.6-70.0) | 342 |
| Foot length (<7.2 cm) | 27 | 45 | 2 | 268 | 37.5 (27.2-49.0) | 99.3 (97.3-99.8) | 93.1 (78.0-98.1) | 85.6 (81.3-89.1) | 50.6 (12.3-207.9) | 0.6 (0.5-0.8) | 68.4 (63.2-73.3) | 342 |
| Foot length (<7.3 cm) | 28 | 44 | 3 | 267 | 38.9 (28.5-50.4) | 98.9 (96.8-99.6) | 90.3 (75.1-96.7) | 85.9 (81.5-89.3) | 35.0 (11.0-111.9) | 0.6 (0.5-0.7) | 68.9 (63.8-73.9) | 342 |
| Foot length (<7.4 cm) | 37 | 35 | 5 | 265 | 51.4 (40.1-62.6) | 98.1 (95.7-99.2) | 88.1 (75.0-94.8) | 88.3 (84.2-91.5) | 27.8 (11.3-68.0) | 0.5 (0.4-0.6) | 74.8 (69.9-79.4) | 342 |
| Foot length (<7.5 cm) | 40 | 32 | 15 | 255 | 55.6 (44.1-66.5) | 94.4 (91.0-96.6) | 72.7 (59.8-82.7) | 88.9 (84.7-92.0) | 10.0 (5.9-17.0) | 0.5 (0.4-0.6) | 75.0 (70.2-79.6) | 342 |
| Foot length (<7.6 cm) | 54 | 18 | 64 | 206 | 75.0 (63.9-83.6) | 76.3 (70.9-81.0) | 45.8 (37.0-54.7) | 92.0 (87.7-94.9) | 3.2 (2.5-4.1) | 0.3 (0.2-0.5) | 75.6 (70.8-80.2) | 342 |
| Foot length (<7.7 cm) | 61 | 11 | 82 | 188 | 84.7 (74.7-91.2) | 69.6 (63.9-74.8) | 42.7 (34.8-50.9) | 94.5 (90.4-96.9) | 2.8 (2.3-3.4) | 0.2 (0.1-0.4) | 77.2 (72.4-81.5) | 342 |
| Foot length (<7.8 cm) | 61 | 11 | 83 | 187 | 84.7 (74.7-91.2) | 69.3 (63.5-74.5) | 42.4 (34.6-50.5) | 94.4 (90.3-96.9) | 2.8 (2.2-3.4) | 0.2 (0.1-0.4) | 77.0 (72.1-81.3) | 342 |
| Foot length (<7.9 cm) | 66 | 6 | 123 | 147 | 91.7 (83.0-96.1) | 54.4 (48.5-60.3) | 34.9 (28.5-42.0) | 96.1 (91.7-98.2) | 2.0 (1.7-2.3) | 0.2 (0.1-0.3) | 73.1 (68.1-77.7) | 342 |
| Foot length (<8 cm) | 69 | 3 | 144 | 126 | 95.8 (88.5-98.6) | 46.7 (40.8-52.6) | 32.4 (26.5-38.9) | 97.7 (93.4-99.2) | 1.8 (1.6-2.0) | 0.1 (0.0-0.3) | 71.3 (66.2-76.1) | 342 |
| Foot length (<8.1 cm) | 71 | 1 | 193 | 77 | 98.6 (92.5-99.8) | 28.5 (23.5-34.2) | 26.9 (21.9-32.5) | 98.7 (93.1-99.8) | 1.4 (1.3-1.5) | 0.0 (0.0-0.3) | 63.6 (58.1-68.6) | 342 |
| Foot length (<8.2 cm) | 71 | 1 | 221 | 49 | 98.6 (92.5-99.8) | 18.1 (14.0-23.2) | 24.3 (19.7-29.5) | 98.0 (89.5-99.6) | 1.2 (1.1-1.3) | 0.1 (0.0-0.5) | 58.4 (53.1-63.8) | 342 |
| Foot length (<8.3 cm) | 71 | 1 | 221 | 49 | 98.6 (92.5-99.8) | 18.1 (14.0-23.2) | 24.3 (19.7-29.5) | 98.0 (89.5-99.6) | 1.2 (1.1-1.3) | 0.1 (0.0-0.5) | 58.4 (53.1-63.8) | 342 |
| Foot length (<8.4 cm) | 72 | 0 | 246 | 24 | 100.0 (94.9-100.0) | 8.9 (6.0-12.9) | 22.6 (18.4-27.6) | 100.0 (86.2-100.0) | 1.1 (1.1-1.1) | 0.0 (.-.) | 54.4 (48.9-59.8) | 342 |
| Foot length (<8.5 cm) | 72 | 0 | 246 | 24 | 100.0 (94.9-100.0) | 8.9 (6.0-12.9) | 22.6 (18.4-27.6) | 100.0 (86.2-100.0) | 1.1 (1.1-1.1) | 0.0 (.-.) | 54.4 (48.9-59.8) | 342 |
| Foot length (<8.6 cm) | 72 | 0 | 261 | 9 | 100.0 (94.9-100.0) | 3.3 (1.8-6.2) | 21.6 (17.5-26.4) | 100.0 (70.1-100.0) | 1.0 (1.0-1.1) | 0.0 (.-.) | 51.7 (46.3-57.2) | 342 |
| Foot length (<8.7 cm) | 72 | 0 | 265 | 5 | 100.0 (94.9-100.0) | 1.9 (0.8-4.3) | 21.4 (17.3-26.1) | 100.0 (56.6-100.0) | 1.0 (1.0-1.0) | 0.0 (.-.) | 50.9 (45.4-56.3) | 342 |
| Foot length (<8.8 cm) | 72 | 0 | 266 | 4 | 100.0 (94.9-100.0) | 1.5 (0.6-3.7) | 21.3 (17.3-26.0) | 100.0 (51.0-100.0) | 1.0 (1.0-1.0) | 0.0 (.-.) | 50.7 (45.4-56.3) | 342 |
| Foot length (<8.9 cm) | 72 | 0 | 270 | 0 | 100.0 (94.9-100.0) | 0.0 (0.0-1.4) | 21.1 (17.1-25.7) |  | 1.0 (1.0-1.0) | .. | 50.0 (44.6-55.4) | 342 |

### S2 Table. Receiver operating curve statistics for accuracy of newborn foot length to classify preterm birth, 1 mm increments (N=342)

| **Preterm birth (<37 weeks vs ≥37 weeks)** | **True positive** | **False negative** | **False positive** | **True negative** | **Sensitivity, % (95% CI)** | **Specificity, % (95% CI)** | **Positive PV, % (95% CI)** | **Negative PV, % (95% CI)** | **Positive LHR, % (95% CI)** | **Negative LHR, % (95% CI)** | **AUC, % (95% CI)** | **Total, N** |
| --- | --- | --- | --- | --- | --- | --- | --- | --- | --- | --- | --- | --- |
| Researcher, overall |  |  |  |  |  |  |  |  |  |  | 85.6 (81.5-89.2) | 342 |
| Foot length (<5.8 cm) | 0 | 25 | 0 | 317 | 0.0 (0.0-13.3) | 100.0 (98.8-100.0) | .. | 92.7 (89.4-95.0) | .. | 1.0 (1.0-1.0) | 50.0 (44.6-55.4) | 342 |
| Foot length (<5.9 cm) | 1 | 24 | 0 | 317 | 4.0 (0.7-19.5) | 100.0 (98.8-100.0) | 100.0 (20.7-100.0) | 93.0 (89.7-95.2) | .. | 1.0 (0.9-1.0) | 52.0 (46.6-57.4) | 342 |
| Foot length (<6 cm) | 1 | 24 | 0 | 317 | 4.0 (0.7-19.5) | 100.0 (98.8-100.0) | 100.0 (20.7-100.0) | 93.0 (89.7-95.2) | .. | 1.0 (0.9-1.0) | 52.0 (46.6-57.4) | 342 |
| Foot length (<6.1 cm) | 2 | 23 | 0 | 317 | 8.0 (2.2-25.0) | 100.0 (98.8-100.0) | 100.0 (34.2-100.0) | 93.2 (90.1-95.5) | .. | 0.9 (0.8-1.0) | 54.0 (48.7-59.5) | 342 |
| Foot length (<6.2 cm) | 2 | 23 | 0 | 317 | 8.0 (2.2-25.0) | 100.0 (98.8-100.0) | 100.0 (34.2-100.0) | 93.2 (90.1-95.5) | .. | 0.9 (0.8-1.0) | 54.0 (48.7-59.5) | 342 |
| Foot length (<6.3 cm) | 2 | 23 | 0 | 317 | 8.0 (2.2-25.0) | 100.0 (98.8-100.0) | 100.0 (34.2-100.0) | 93.2 (90.1-95.5) | .. | 0.9 (0.8-1.0) | 54.0 (48.7-59.5) | 342 |
| Foot length (<6.4 cm) | 2 | 23 | 0 | 317 | 8.0 (2.2-25.0) | 100.0 (98.8-100.0) | 100.0 (34.2-100.0) | 93.2 (90.1-95.5) | .. | 0.9 (0.8-1.0) | 54.0 (48.7-59.5) | 342 |
| Foot length (<6.5 cm) | 3 | 22 | 0 | 317 | 12.0 (4.2-30.0) | 100.0 (98.8-100.0) | 100.0 (43.8-100.0) | 93.5 (90.4-95.7) | .. | 0.9 (0.8-1.0) | 56.0 (50.7-61.5) | 342 |
| Foot length (<6.6 cm) | 3 | 22 | 4 | 313 | 12.0 (4.2-30.0) | 98.7 (96.8-99.5) | 42.9 (15.8-75.0) | 93.4 (90.3-95.6) | 9.5 (2.3-40.2) | 0.9 (0.8-1.0) | 55.4 (49.8-60.6) | 342 |
| Foot length (<6.7 cm) | 3 | 22 | 4 | 313 | 12.0 (4.2-30.0) | 98.7 (96.8-99.5) | 42.9 (15.8-75.0) | 93.4 (90.3-95.6) | 9.5 (2.3-40.2) | 0.9 (0.8-1.0) | 55.4 (49.8-60.6) | 342 |
| Foot length (<6.8 cm) | 3 | 22 | 4 | 313 | 12.0 (4.2-30.0) | 98.7 (96.8-99.5) | 42.9 (15.8-75.0) | 93.4 (90.3-95.6) | 9.5 (2.3-40.2) | 0.9 (0.8-1.0) | 55.4 (49.8-60.6) | 342 |
| Foot length (<6.9 cm) | 5 | 20 | 6 | 311 | 20.0 (8.9-39.1) | 98.1 (95.9-99.1) | 45.5 (21.3-72.0) | 94.0 (90.9-96.1) | 10.6 (3.5-32.2) | 0.8 (0.7-1.0) | 59.1 (53.6-64.3) | 342 |
| Foot length (<7 cm) | 7 | 18 | 8 | 309 | 28.0 (14.3-47.6) | 97.5 (95.1-98.7) | 46.7 (24.8-69.9) | 94.5 (91.5-96.5) | 11.1 (4.4-28.1) | 0.7 (0.6-0.9) | 62.7 (57.5-68.0) | 342 |
| Foot length (<7.1 cm) | 10 | 15 | 14 | 303 | 40.0 (23.4-59.3) | 95.6 (92.7-97.4) | 41.7 (24.5-61.2) | 95.3 (92.4-97.1) | 9.1 (4.5-18.3) | 0.6 (0.5-0.9) | 67.8 (62.6-72.8) | 342 |
| Foot length (<7.2 cm) | 14 | 11 | 15 | 302 | 56.0 (37.1-73.3) | 95.3 (92.3-97.1) | 48.3 (31.4-65.6) | 96.5 (93.8-98.0) | 11.8 (6.5-21.6) | 0.5 (0.3-0.7) | 75.6 (70.8-80.2) | 342 |
| Foot length (<7.3 cm) | 15 | 10 | 16 | 301 | 60.0 (40.7-76.6) | 95.0 (92.0-96.9) | 48.4 (32.0-65.2) | 96.8 (94.2-98.2) | 11.9 (6.7-21.1) | 0.4 (0.3-0.7) | 77.5 (72.7-81.8) | 342 |
| Foot length (<7.4 cm) | 17 | 8 | 25 | 292 | 68.0 (48.4-82.8) | 92.1 (88.6-94.6) | 40.5 (27.0-55.5) | 97.3 (94.8-98.6) | 8.6 (5.4-13.7) | 0.3 (0.2-0.6) | 80.1 (75.5-84.2) | 342 |
| Foot length (<7.5 cm) | 17 | 8 | 38 | 279 | 68.0 (48.4-82.8) | 88.0 (84.0-91.1) | 30.9 (20.3-44.0) | 97.2 (94.6-98.6) | 5.7 (3.8-8.5) | 0.4 (0.2-0.6) | 78.0 (73.3-82.3) | 342 |
| Foot length (<7.6 cm) | 20 | 5 | 98 | 219 | 80.0 (60.9-91.1) | 69.1 (63.8-73.9) | 16.9 (11.2-24.7) | 97.8 (94.9-99.0) | 2.6 (2.0-3.3) | 0.3 (0.1-0.6) | 74.5 (69.6-79.1) | 342 |
| Foot length (<7.7 cm) | 22 | 3 | 121 | 196 | 88.0 (70.0-95.8) | 61.8 (56.4-67.0) | 15.4 (10.4-22.2) | 98.5 (95.7-99.5) | 2.3 (1.9-2.8) | 0.2 (0.1-0.6) | 74.9 (69.9-79.4) | 342 |
| Foot length (<7.8 cm) | 22 | 3 | 122 | 195 | 88.0 (70.0-95.8) | 61.5 (56.1-66.7) | 15.3 (10.3-22.0) | 98.5 (95.6-99.5) | 2.3 (1.9-2.8) | 0.2 (0.1-0.6) | 74.8 (69.9-79.4) | 342 |
| Foot length (<7.9 cm) | 24 | 1 | 165 | 152 | 96.0 (80.5-99.3) | 47.9 (42.5-53.4) | 12.7 (8.7-18.2) | 99.3 (96.4-99.9) | 1.8 (1.6-2.1) | 0.1 (0.0-0.6) | 72.0 (66.8-76.6) | 342 |
| Foot length (<8 cm) | 24 | 1 | 189 | 128 | 96.0 (80.5-99.3) | 40.4 (35.1-45.9) | 11.3 (7.7-16.2) | 99.2 (95.7-99.9) | 1.6 (1.4-1.8) | 0.1 (0.0-0.7) | 68.2 (62.9-73.0) | 342 |
| Foot length (<8.1 cm) | 24 | 1 | 240 | 77 | 96.0 (80.5-99.3) | 24.3 (19.9-29.3) | 9.1 (6.2-13.2) | 98.7 (93.1-99.8) | 1.3 (1.1-1.4) | 0.2 (0.0-1.1) | 60.1 (54.8-65.5) | 342 |
| Foot length (<8.2 cm) | 24 | 1 | 268 | 49 | 96.0 (80.5-99.3) | 15.5 (11.9-19.8) | 8.2 (5.6-11.9) | 98.0 (89.5-99.6) | 1.1 (1.0-1.2) | 0.3 (0.0-1.8) | 55.7 (50.4-61.2) | 342 |
| Foot length (<8.3 cm) | 24 | 1 | 268 | 49 | 96.0 (80.5-99.3) | 15.5 (11.9-19.8) | 8.2 (5.6-11.9) | 98.0 (89.5-99.6) | 1.1 (1.0-1.2) | 0.3 (0.0-1.8) | 55.7 (50.4-61.2) | 342 |
| Foot length (<8.4 cm) | 24 | 1 | 294 | 23 | 96.0 (80.5-99.3) | 7.3 (4.9-10.7) | 7.5 (5.1-11.0) | 95.8 (79.8-99.3) | 1.0 (1.0-1.1) | 0.6 (0.1-3.9) | 51.6 (46.3-57.2) | 342 |
| Foot length (<8.5 cm) | 24 | 1 | 294 | 23 | 96.0 (80.5-99.3) | 7.3 (4.9-10.7) | 7.5 (5.1-11.0) | 95.8 (79.8-99.3) | 1.0 (1.0-1.1) | 0.6 (0.1-3.9) | 51.6 (46.3-57.2) | 342 |
| Foot length (<8.6 cm) | 25 | 0 | 308 | 9 | 100.0 (86.7-100.0) | 2.8 (1.5-5.3) | 7.5 (5.1-10.8) | 100.0 (70.1-100.0) | 1.0 (1.0-1.0) | 0.0 (.-.) | 51.4 (46.0-56.9) | 342 |
| Foot length (<8.7 cm) | 25 | 0 | 312 | 5 | 100.0 (86.7-100.0) | 1.6 (0.7-3.6) | 7.4 (5.1-10.7) | 100.0 (56.6-100.0) | 1.0 (1.0-1.0) | 0.0 (.-.) | 50.8 (45.4-56.3) | 342 |
| Foot length (<8.8 cm) | 25 | 0 | 313 | 4 | 100.0 (86.7-100.0) | 1.3 (0.5-3.2) | 7.4 (5.1-10.7) | 100.0 (51.0-100.0) | 1.0 (1.0-1.0) | 0.0 (.-.) | 50.6 (45.2-56.0) | 342 |
| Foot length (<8.9 cm) | 25 | 0 | 317 | 0 | 100.0 (86.7-100.0) | 0.0 (0.0-1.2) | 7.3 (5.0-10.6) | .. | 1.0 (1.0-1.0) | .. | 50.0 (44.6-55.4) | 342 |
